# Increasing frequency of hospital admissions for retinal detachment and vitreo-retinal surgery in England 2000-2018

**DOI:** 10.1101/2020.11.06.20214734

**Authors:** Haifa A Madi, Johannes Keller

## Abstract

**Objectives:** To analyse the changes in reported frequency of retinal detachment admissions and vitreo-retinal surgery procedures performed between 2000-2018 in England. To obtain information useful to contribute towards the planning of service delivery.

**Methods:** Analysis of England’s Hospital Episode Statistics from the Health and Social Care Information Centre and population data from the United Kingdom’s Office for National Statistics.

**Results:** Episodes of “retinal detachments with breaks” increased year on year from 3,447 (7.0/100M) in 2000 to 10,971 (19.7/100M) in 2018 (p<0.001), whereas records of “tractional retinal detachment” increased from 290 (0.6/100M) to 910 (1.6/100M) in the same period (p<0.0001). The number of reported pars plana vitrectomies irrespective of indication increased over fourfold from 5,761 to 26,900 (p<0.0001), while the number of scleral buckling records decreased by two thirds from 2,897 to 780 (p<0.0001). During the same period the population of England increased from 49.2 million-55.6 million, proportionally at a slower rate than that for recorded hospital episodes.

**Conclusions:** The frequency of admissions to hospital for surgically treated retinal detachment seems to have been increasing significantly since 2000. This effect is more marked in cases of rhegmatogenous retinal detachment. This may be partially explained by repeat surgery in cases of recurrent retinal detachment. Other possible explanations may be increased incidence of disease (due to increased rates of cataract surgery, increasing longevity and rates of myopia), improvements in patient access, and increased public awareness. It is possible that this observation is due in part to local changes in methodology of hospital coding.

## Introduction

Rhegmatogenous retinal detachment (RRD) is a serious ophthalmic condition where the neurosensory retina becomes separated from the retinal pigment epithelium through ingress of fluid from a retinal break. Without treatment, RRD leads to blindness in the affected eye. Advances in surgical treatment allow for the prevention of vision loss in most cases. Recent refinements in surgical technique including self-sealing small gauge vitrectomy and wide-angle viewing systems have significantly improved the ability of surgeons to treat RRD quickly and safely.

In the United Kingdom, the annual incidence of RRD is reported to have increased from 6.1-9.8 cases per 100,000 people during the 1970’s, to 11.8-17.9 cases per 100 000 people in the 1990s^1^. A more recent report estimated an incidence increase from 13.4/100,000 in the 1990s to 15.4/100,000 in the 2010s^2^. A meta-analysis of 5 studies carried out in Europe^3^ estimated the average incidence of RD in the continent at 13.3/100,000.

In England, all instances of hospital admission, day case intervention and emergency attendances are recorded in a database containing details of diagnoses and procedures called the Hospital Episode Statistics (HES)^4^. Each instance of care is recorded as one episode, regardless if a given patient is treated more than once for the same condition. Diagnoses are recorded using the World Health Organisation International Classifications of Diseases version 10 (ICD-10) classification system and procedures are coded with the NHS’s Office for Population Census and Surveys Classification of Surgical Operations version 4 (OPCS-4). These data are identified and coded from medical records following the discharge of each patient from each episode. English NHS healthcare providers use these data to claim refund from the regional funding bodies for the care they deliver. The same data are also processed and used for other purposes, such as health services delivery planning and research.

We set out to analyse England’s HES to seek changes in trends of admission to healthcare facilities for treatment of RRD from 2000 to 2018. We also set out to study changes in frequency and type of vitreo-retinal operations carried out during this period. The purpose was to gain insight into the epidemiology of surgical retinal disease with the aim to inform service planning and resource deployment.

## Materials and Methods

The publicly available HES Hospital Admitted Patient Care Activity records were downloaded from the NHS Digital^4^ internet portal both for procedures and diagnosis. The reported year follows the UK fiscal year which runs from April until March and is reported based on the year of period end, therefore 2004 comprises the period from April 2003 until March 2004. England’s population data were obtained from the mid-year estimate from the Office for National Statistics^5^ corresponding to the fiscal year where the estimate lies. For example, the mid-year estimate for 2010 was used to calculate the rate of admission for the fiscal year 2010-11.

### Diagnoses

The ICD-10 codes for retinal detachment with breaks (H33.0) and tractional retinal detachment (TRD) (H33.4) were analysed separately. The code H33.0 explicitly excludes cases where the retinal detachment was associated with diabetes which should allow for a targeted analysis of RRD. There is no similar distinction within code H33.4 between diabetes-related tractional detachment and other tractional pathology such as retinopathy of prematurity or proliferative vitreo-retinopathy. Associated codes within the same H33.x root include codes for retinoschisis and retinal cysts H33.1, serous retinal detachment H33.2, retinal breaks without detachment H33.3. We analysed changes in H33.2 and H33.3 as comparators as they should be susceptible to variation due to population change. The code for other retinal detachments H33.5 was not analysed.

### Procedures

The OPCS-4 codes for interventions on the retina and the lens were studied. Records were analysed from the database of main procedure descriptor for pars plana vitrectomy (C79.2) and cataract surgery using insertion of prosthetic replacement for lens (C75.1) as recommended by the NHS Classification Service. For scleral buckle operations we aggregated all codes describing the implantation of a scleral indenting device or otherwise creating scleral indent on the retina: overlay scleroplasty (C54.1), imbrication of sclera (C54.2), buckling of sclera and implant (C54.3), buckling of sclera and local or encircling explant (C54.4), buckling of sclera (C54.5), other specified buckling operations for attachment of retina (54.8) and unspecified buckling operations for attachment of retina (54.9).

Additionally, as there is no nationally mandated coding record of pars plana vitrectomy in primary position for all vitrectomy operations, we built a construct adding surgical manoeuvres which require pars plana vitrectomy access to the vitreous cavity, to the vitrectomy code (C79.2). Therefore the following codes were grouped with C79.2 and analysed as a construct: injection of vitreous substitute into vitreous body NEC (C79.3), internal tamponade of retina using liquid (C79.6), removal of internal tamponade agent from vitreous body (C79.7), other specified operations on vitreous body (C79.8), peel of epiretinal fibroglial membrane (C80.1), peel of internal limiting membrane (C80.2), delamination of epiretinal fibrovascular membrane (C80.3), segmentation of epiretinal fibrovascular membrane (C80.4), removal of subretinal vascular membrane (C80.5), removal of subretinal membrane NEC (C80.6), other specified operations on retinal membrane (C80.8) unspecified operations on retinal membrane (C80.9), pigment epithelium translocation of retina (C83.1), macular translocation three hundred and sixty degrees (C83.2), limited macular translocation (C83.3), other specified translocation of retina (C83.8), unspecified translocation of retina (C83.9), epiretinal dissection (C84.1), excision of lesion of retina NEC (C84.2), biopsy of lesion of retina (C84.3), retinal vascular sheathotomy (C84.4), drainage of subretinal fluid through retina (C84.5), retinotomy NEC (C84.6), other specified other operations on retina (C84.8). As laser therapy can be administered either transpupillarily or with endolaser probes, the codes for laser intervention were not included.

### Statistical Analysis and variable associations

Descriptive and linear regression analysis was carried out for diagnostic and intervention variables. Linear regression analysis was carried out for variables potentially associated with RD including cataract surgery volumes. The ratio between buckling operations and admissions for RRD was calculated.

Consent was not required as this is a study of a database in the public domain and no patient identifiable information was used.

## Results

### Diagnoses

During the business year ending 2001 there were 3,447 admissions to hospitals in England for RRD with breaks (Figure 1). This figure more than trebled with an average annual growth of 7.2% and a slope of 464.7 cases per year to 10,971 in 2018 (R = 0.98, R^2^ = 0.97, p <0.001). TRDs during 2000-01 were recorded in 290 instances. These too increased steadily at an annual rate of 7.9%, more than trebling in the study period to 910 (R=0.93, R2=0.87, p=<0.0001). By comparison, serous retinal detachments were recorded with a frequency of 3,941 in 2000 but reduced slightly throughout the study period with a regression slope of −32.1 (R=−0.56, R^2^ = 0.32, p=0.01). Retinal breaks without a detachment were recorded at 3,141 in 2000, dropping to 1,842 in 2004 and then remaining relatively stable from 2006 with an average of 2433.1 per annum (R=−0.25, R2=0.06, p=0.93).

**Figure 1:**
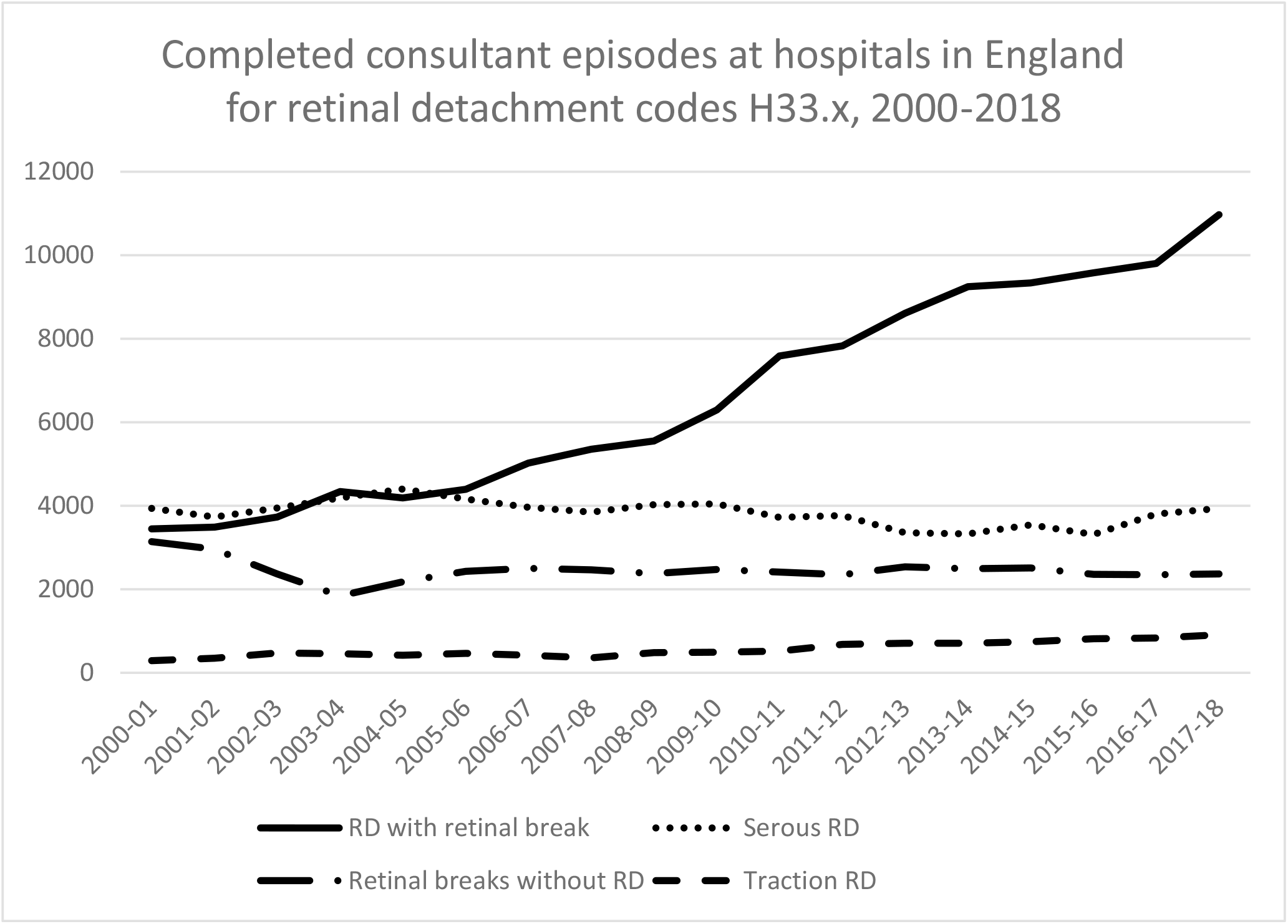
Completed consultant episodes at hospitals in England for retinal detachment codes H33.x, 2000-2018.

Calculating the rate of admission for the mid-year population level showed 7.0 episodes per 100,000 population for RD with breaks during 2000-01 with year-on-year increases to 19.7/100,000 in 2017-18 with a slope of 0.79 and R=0.98. The slopes for admissions for TRD (0.05/year), serous retinal detachment (−0.12/year) and retinal breaks without detachment (−0.06/year) show little change compared to population growth.

To estimate the effect of re-admission for recurrent retinal detachment, we assumed an average RRD recurrence rate of 13.0%^6^and a single operation to repair the recurrence. Deducting these from the total of admissions to estimate the value of primary retinal detachments, the regression slope is reduced to 404.3 cases per annum of growth (Figure 2). Assuming 2.5 operations to repair a recurrent retinal detachment further lowers the slope to 312.6 from 2,318.1 cases in 2000 to 7,378.0 cases in 2018.

**Figure 2:**
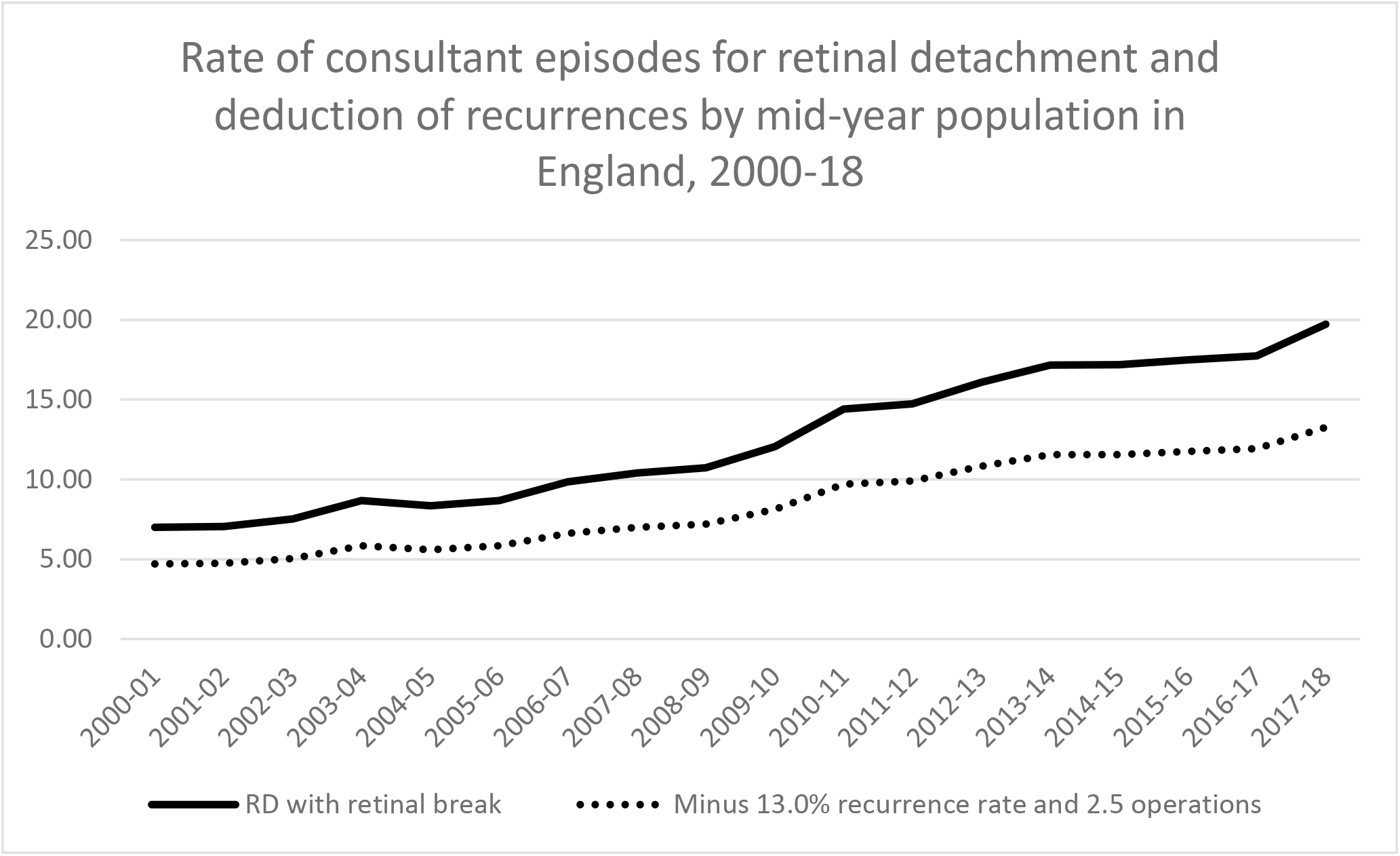
Hospital admissions for retinal detachment and deduction of RD recurrence estimate by mid-year population in England 2000-2018.

### Procedures

During 2000-01 there were 5,761 pars plana vitrectomy procedures carried out and recorded as main intervention. Adding the codes of other surgical manoeuvres recorded as main intervention and which obligatorily required pars plana vitrectomy, the total estimate of pars plana vitrectomy rises to 8,777. Both figures have increased steadily with a yearly rate of 8.5% for vitrectomy (slope = 1,384.1, R^2^=0.97, p<0.0001) and 6.9% for the composite of vitrectomy codes (slope=1,363.3, R^2^=0.99, p<0.0001). The final figure for 2017-18 is 26,900 for vitrectomy coded singly and 29,923 for vitrectomy composite respectively (Figure 3). Similarly, for 2000-01 there were 2,897 scleral buckling operations which decreased steadily at an average yearly rate of 7.0% to 780 cases with a slope of - 114.5 (R^2^=0.87, p<0.0001)

**Figure 3:**
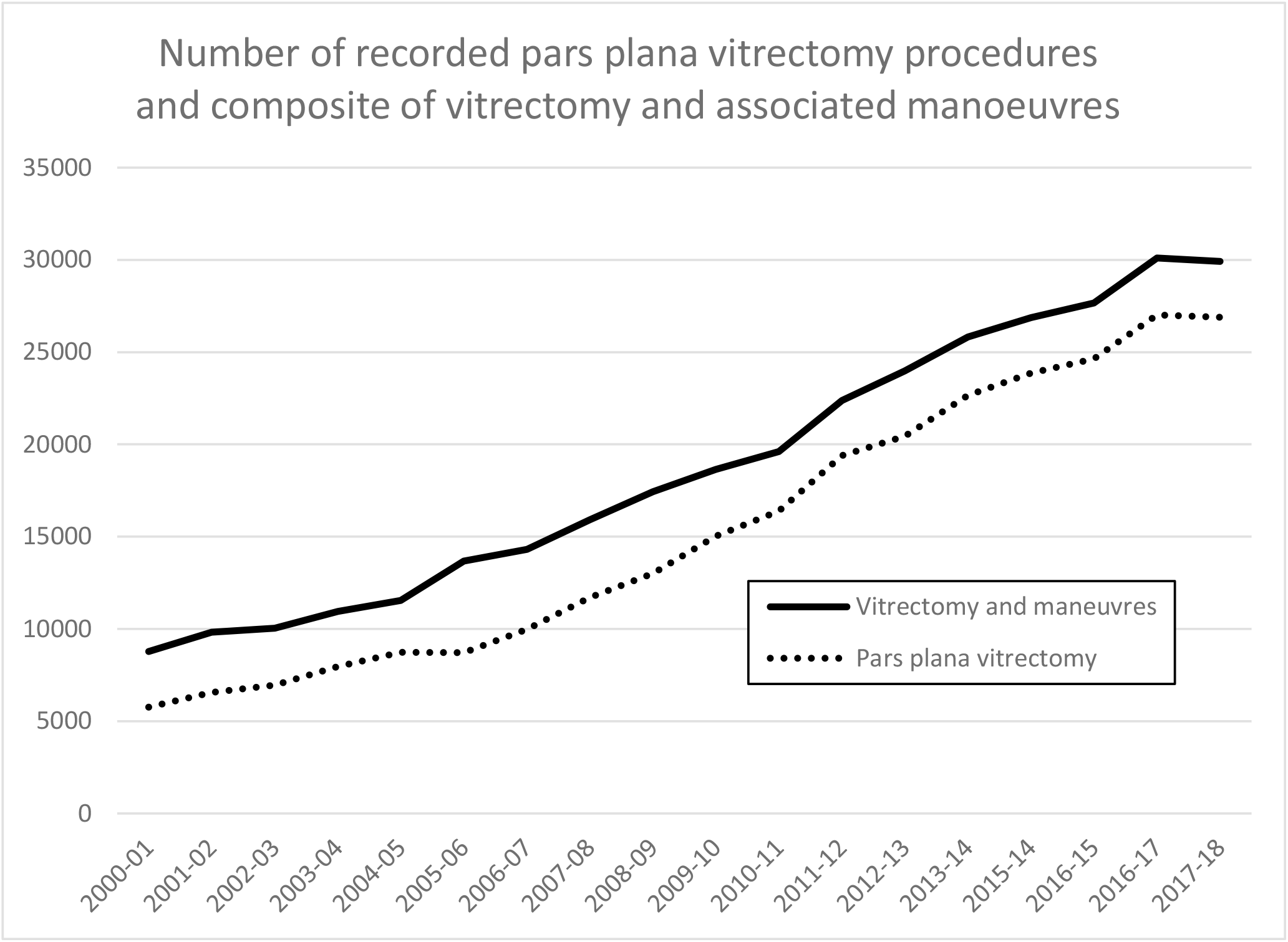
Number of recorded pars plana vitrectomy procedures and composite of vitrectomy and associated manoeuvres.

**Figure 4:**
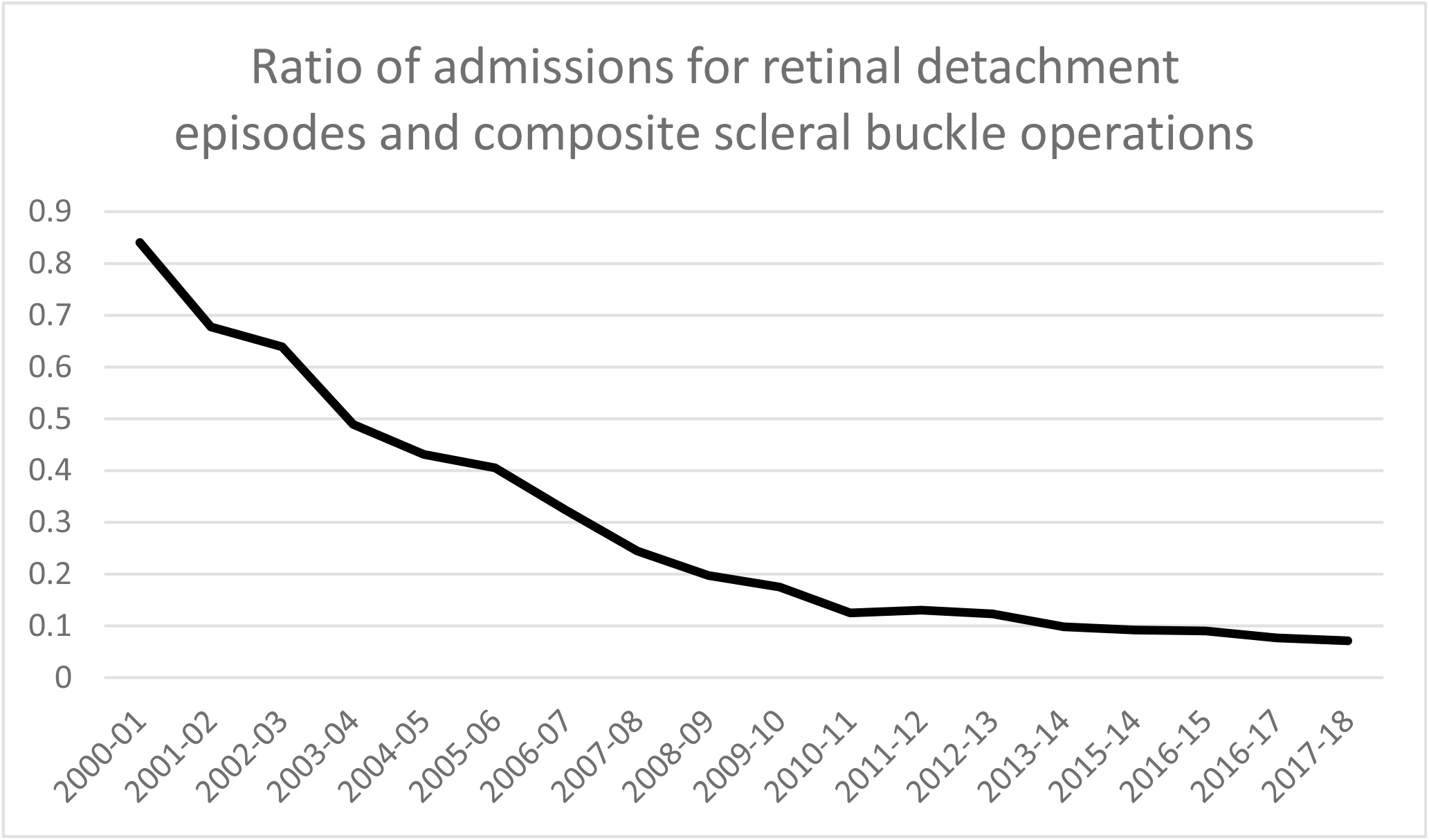
Ratio of admissions for retinal detachment episodes and composite scleral buckle operations.

### Associations

The ratio of scleral buckle operations to RRD admissions changed from 84:100 in 2000-01 to 7.1:100 in 2017-18, at a rate of −0.04 buckling operation per admission for RRD per year. There was a strong and statistically significant correlation between the number of cataract surgery procedures and number of admissions for RRD with breaks (r=0.932, r2=0.868, p<0.0001).

## Discussion

We found that the frequency of admissions to hospital for the types of retinal detachment which are treated by surgery (both TRD and RRD) increased steadily in England from 2000-2018. During the same period, England’s population increased from 49.2million to 54.3million, at a proportionally slower rate than that for recorded hospital episodes. It is important to make the distinction between incidence, prevalence and frequency of a disease. As the HES collect episodes, it is not possible to determine neither incidence nor prevalence without obtaining the raw HES data to eliminate duplicates as any episode of care for the same condition will be counted twice. However, given that the methodology of data collection for HES can be assumed to be reasonably constant, trend analysis for defined variables is useful in gaining understanding of the behaviour of healthcare activity.

### Rhegmatogenous retinal detachment

There have been several national and regional population studies published over recent years which have estimated the annual incidence of RRD, reporting results ranging from 9.7 to 20.7/100,000/year in developed Western and Asian nations^1-2,7-14^. Broadly, two types of methodology were used: hospital records analysis and national reimbursement databases. Hospital records analysis have been reported for defined populations where only a small number of hospitals provide retinal detachment care. Manners^7^ analysed all admissions to Western Australian hospitals between 2000-2013 reporting stable rates between 12.8 and 16.2/100,000 people. Also in Australia, Howie^8^ reported rates for the island-state of Tasmania where a single surgeon provided care between 2005 and 2010, finding a rate of 9.7/100,000 inhabitants. In Scotland^9^, a consortium formed by 6 hospitals prospectively recorded all primary cases between 2007-2009 finding an incidence of 12.05/100,000. A similar consortium formed in the Netherlands^10^ reported 18.2/100,000 cases during 2009. A further report from a single centre in the city of Odense in Denmark^11^estimated an incidence of 20.7/100,000 however being a tertiary referral centre, it is possible that it also treated patients from outside its reference area. This type of study is better suited to define clinical variables and patient characteristics.

Analysis of national databases have also been reported studying recent years in England^1,2^, Denmark^12^, and Korea^13^. These are more likely to give more accurate estimates and allow to describe time-trends over a period. In England, multiple database linkage analysis was carried out covering the period from 1968 through to 2011. This study also sought to find associations with socioeconomic and health status variables. Analysing the general three-digit code for retinal detachment H33 without distinction, it estimated that the global incidence of all types of retinal detachments increased from 13.4/100,000 in 1999 to 15.4/100,000 in 2011. A similar nationwide study in Denmark^12^ also found a steady increase of incidence from 2000 to 2011 which they linked to an increase in rates of cataract surgery. The mean incidence over the period was reported at 13.7/100,000. In Korea^13^, a five-year study covering 2007 to 2011 found an incidence of 10.4/100,000 which remained stable. A further Taiwanese^14^ database covering 4% of the population reported an estimated incidence of 16.4/100,000 cases from 2000-2012.

It is quite remarkable that in a period of 18 years, the rate for admissions for RRD in England went from 7.0/100,000 to 19.7/100,000 people. Even when including an estimate for recurrent cases, this represents a steep increase which almost doubles the figure of admissions to 13.7/100,000.

Several risk factors predisposing to RRD have been identified including male gender, increasing age, previous cataract surgery, and myopia. Several of the database studies cited above have found that age is an important risk factor for RRD^2,12,14^. Shah found an almost 20-fold increased risk of RD in the over 60s age group compared with the under 30s age group^2^. As the average age of England’s population increases, so has the average age of admissions for RRD. While male gender is a significant risk factor for RRD, there have been no recent significant changes in the gender composition of England’s population^5^. There are no accurate population-wide studies of the prevalence of myopia in England. However, analysis of population cohorts has shown a sustained worldwide increase in the prevalence of myopia particularly amongst young subjects. This effect is more marked across Asia, Europe and the United States^15^. An eye with a refractive error of more than −3 dioptres has a tenfold increased risk of RD compared to an emmetropic eye^9^. The increase in the number of performed cataract operations can be partly attributed to population aging, and hence a higher prevalence of cataracts. In addition, there has been an inclination to perform cataract surgery at an earlier stage with the advent of phacoemulsification^10^. A review of the literature found that the risk of RD following phacoemulsification is approximately ten times the risk for the general population^16^.

It is interesting to note that the rate of retinal breaks remained relatively constant during the period analysed. This observation is surprising because by definition RRDs occur after retinal tear formation. Therefore if the natural rate of RD was increasing, it could be expected that the rate of retinal tears would also have increase in parallel or faster. A potential explanation for this observation is a proposed catastrophic behaviour of RRD development, where they would form suddenly in “leaps and bounds”^17^, therefore only low-risk tears would be diagnosed prior to developing into RD. Another potential explanation would be improvements in access to care for patients both by improved referral pathways and by offering surgery to RDs which previously would have been deemed inoperable. Further, we observed that when a correction was introduced to account for estimated rates of recurrent RD, the slope of the growth in RD episodes became less steep. We believe that it is likely that improved instrumentation has led to practitioners persevering in achieving retinal attachment through multiple operations. Operating on recurrent RD would then partially explain the marked increase in HES recorded episodes. It is worth noting that regardless of the incidence of primary RD, a proportion of patients will unfortunately experience RD recurrences. This fact needs to be considered when planning the delivery of service.

### Tractional retinal detachment

In our study we also observed a big increase in TRD admissions. The prevalence of diabetes mellitus increased year on year since 2005-2018 from 3.6%-5.8%, which can partly account for the trend we observed^18^. However a group of researchers^19^ found reducing rates of vitrectomy for all indications in diabetic patients in a well-defined geographic area in the North-East of England. They attributed this to better screening and early treatment of proliferative diabetic retinopathy. In recent years, public health intervention on diabetic eye disease in England has been robust, likely reducing the proportion of patients developing severe disease. Therefore as the likelihood of developing TRD for any given patient is decreasing, the number of new diabetics is increasing at a large rate. Additionally, the code for TRD allows for coding cases caused by proliferative vitreo-retinopathy, retinopathy of prematurity and rarer forms of TRD, making it more difficult to analyse diabetes cases in isolation. Another factor which may contribute to explain this increase are improvements in surgical techniques which have resulted in lower thresholds for intervention.

### Increase in vitrectomy

We found that the number of reported pars plana vitrectomies irrespective of indication increased over fourfold between 2000-2018. When analysing data since the introduction of vitrectomy surgery, the rate of increase in England was near tenfold up to 2003^1^. Advances in surgical instrumentation, massification of the technology, the introduction of optical coherence tomography have all contributed to the expanding application of vitrectomy surgery to a larger number of conditions. Unfortunately, as ICD-10 codes for macular disease make no distinction between conditions treated medically or surgically, analysis of the data on HES does not help to elucidate changes in rates of macular surgery.

### Decrease in scleral buckling

Once the mainstay treatment for RD, the number of scleral buckling performed decreased by two thirds. This observation was already apparent in the data analysed up to 2004^1^. This change coincides with significant improvements in operating microscopes and surgical instrumentation, as well as important advances in surgical techniques and training. A landmark randomised controlled trial^20^ published in 2007 showed no significant difference in the likelihood of achieving retinal attachment overall except in pseudophakic eyes where it found an advantage for vitrectomy. Despite scleral buckling allowing better visual outcomes in phakic eyes, vitrectomies have overwhelmingly become the preferred surgical strategy for RRD. Presumably the effectiveness, safety and speed of the operation have contributed to this. This poses challenges for training as a reduced frequency of buckling surgery will potentially undermine the success rate of the operation which remains the state of the art for certain types of RRD. Encouragingly, the ratio of scleral buckling seems to have stabilised from 2015.

### Limitations of this study

The use of national datasets has the advantage of providing large scale data over a long time span. However, they are not without their limitations^21^. For one, registry studies offer no clinical data, such as lens status or axial length, which are significant risk factors for RRD. Secondly, their validity depends on the accuracy with which clinical information is reported and coded. The risk of missing cases due to the lack of coding in this study is presumably low because coding is necessary for reimbursing expenses in the NHS since the introduction of the “payment by result” policy in 1990. However, not all NHS hospitals may be equally adept at obtaining appropriate reimbursement and practices in coding may vary slightly from hospital to hospital. For example, not all hospitals may code “pars plana vitrectomy” as the defining procedure, but another step done during the operation. For this reason, adding OPCS manoeuvres which can only be done as part of a vitrectomy operation gives a higher number, likely to be closer to the true number of vitrectomies carried out. Given that these differences and potential errors in coding are likely to be systematic, HES studies are useful in understanding trends. Another issue with coding is the way TRD is defined, not only used for diabetic related RD but for other aetiologies of TRD. Shah^2^ found that the increase in RD admissions in England was likely to be associated with diabetes through linkage analysis with the Oxford database. There is however no biological plausibility to diabetes leading to an increase in RRD. As the study used the generic three-digit code for RD (H33), rather than breaking it down into subcategories (H33.X). We found that both TRD and RRD increased significantly. It is possible that some RD “with breaks” which should be coded H33.0 may have been coded as H33.4 “tractional RD” and vice-versa, even if the code for RRD specifically excludes diabetes as an aetiology. Another limitation of analysing aggregated HES reports is that as episodes are recorded per admission, data about individual patients cannot be identified. Therefore multiple entries are made for the same patient who has had more than one admission for vitreo-retinal surgery. The reported frequency of RD may be underestimated by failing to record cases who opt not to have surgery. In addition, a small number of patients may have undergone surgery abroad or in private hospitals not supplying data to NHS Digital. Given the make-up of England’s healthcare provision, these numbers are likely to be very small. For all the above reasons, caution must be taken in making clinical inferences from analysing administrative data such as England’s HES.

In conclusion, the hospital admission rates for all types of RD seem to have increased significantly in recent years, an effect more marked in cases of RRD. This observation is probably attributable to increasing age, rise in the number of performed cataract operations and increase in the prevalence of diabetes for the tractional type of retinal detachment. Other possible explanations may be increased prevalence of myopia, changes in methodology of hospital coding, improvements in patient access and public awareness, increased repeated procedures, and lower thresholds for operating on a chronic RRD with poor prognosis. Despite the disruption that Covid-19 has brought to medical care, a good understanding of these trends should allow a more accurate prediction of future demand for vitreo-retinal surgery across England.

## Data Availability

The data is publicly available at NHS Digital. https://digital.nhs.uk/data-and-information/publications/statistical/hospital-admitted-patient-care-activity

